# A master protocol to investigate a novel therapy acetyl-L-leucine for three ultra-rare neurodegenerative diseases: Niemann-Pick type C, the GM2 Gangliosidoses, and Ataxia Telangiectasia

**DOI:** 10.1101/2020.03.11.20034256

**Authors:** T. Fields, M. Patterson, T. Bremova, G. Belcher, I. Billington, G.C. Churchill, W. Davis, W. Evans, S. Flint, A. Galione, U. Granzer, J. Greenfield, R. Karl, R. Kay, D. Lewi, T. Mathieson, T. Meyer, D. Pangonis, F.M. Platt, L. Tsang, C. Verburg, M. Factor, M. Strupp

**Affiliations:** IntraBio Ltd, Begbroke Science Park, Begroke Hill, Woodstock Road, Oxford OX5 1PF, United Kingdom; Mayo Clinic, 200 First Street SW, Rochester MN 55905, United States; Department of Neurology, Inselspital, University Hospital Bern, and University of Bern, Bern, Switzerland; PV Consultancy, 113 St Georges Square Mews, London SW1V 3RZ, United Kingdom; Department of Pharmacology, University of Oxford, Mansfield Road, Oxford OX1 3QT, United Kingdom; Ataxia-Telangiectasia Society, Rothamsted Experimental Station West Common, Harpenden AL5 2JQ, United Kingdom; Niemann-Pick UK, Suite 2, Vermont House, Concord, Washington, Tyne and Wear, NE37 2SQ; Primary Care Stratified Medicine (PRISM), Division of Primary Care, University of Nottingham, Nottingham; Granzer Regulatory Consulting & Services, Kistlerhofstr. 172C, D-81379 München, Germany; Ataxia UK, 12 Broadbent close, London N6 5JW, United Kingdom; Cure Tay-Sachs Foundation 2409 E. Luke Avenue, Phoenix AZ, 85016, Unites States; RK Statistics, Brook House, Mesne Lane, Bakewell DE45 1AL, United Kingdom; The Cure & Action for Tay-Sachs (CATS) Foundation, 94 Milborough Crescent, Lee, London SE12 0RW, United Kingdom; International Niemann-Pick Disease Alliance, Suite 2, Vermont House, Concord, Washington, Tyne and Wear, NE37 2SQ, United Kingdom; National Tay-Sachs and Allied Disease Foundation; 2001 Beacon Street, Suite 204, Boston, MA 02135, United States; Arnold & Porter Kaye Scholer LLP, 25 Old Broad Street | London EC2N 1HQ, United Kingdom; Department of Neurology and German Center for Vertigo and Balance Disorders, Ludwig Maximilians University, Munich, Germany

**Keywords:** Niemann-Pick type C (NPC), GM2 gangliosidosis, Tay-Sachs disease (TSD), Sandhoff disease, ataxia telangiectasia, N-acetyl-L-leucine, pharmaceutical intervention, symptomatic treatment, single-blinded trial, cerebellar ataxia, lysosomal storage disease

## Abstract

**Background:** The lack of approved treatments for the majority of rare diseases is reflective of the unique challenges of orphan drug development. Novel methodologies, including new functionally relevant endpoints, are needed to render the development process more feasible and appropriate for these rare populations, and thereby expedite the approval of promising treatments to address patients’ high unmet medical need. Here, we describe the development of an innovative master protocol and primary outcome assessment to investigate the modified amino acid N-acetyl-L-leucine (Sponsor Code: IB1001) in three seperate, multinational, phase II trials for three ultra-rare, autosomal-recessive, neurodegenerative disorders: Niemann-Pick disease type C (NPC), GM2 Gangliosidoses (Tay-Sachs and Sandhoff disease; “GM2”), and Ataxia Telangiectasia (A-T).

**Methods/design:** The innovative IB1001 master protocol and novel CI-CS primary endpoints were developed through a close collaboration between the Industry Sponsor, Key Opinion Leaders, representatives of the Patient Communities, and National Regulatory Authorities. As a result, the open-label, rater-blinded study design is considerate of the practical limitations of recruitment and retention of subjects in these ultra-orphan populations. The novel primary endpoint, the Clinical Impression of Change in Severity^©^ (CI-CS), accommodates the heterogenous clinical presentation of NPC, GM2, and A-T: at screening, the Principal Investigator appoints for each patient a primary anchor test (either the 8 Meter Walk Test (8MWT) or 9 Hole Peg Test of the Dominant Hand (9HPT-D)) based on his/her unique clinical symptoms. The anchor tests are videoed in a standardized manner at each visit to capture all aspects related to the patient’s functional performance. The CI-CS assessment is ultimately performed by independent, blinded raters who compare videos of the primary anchor test from three periods: baseline, the end of treatment, and the end of a post-treatment washout. Blinded to the timepoint of each video, the raters make an objective comparison scored on a 7-point Likert scale of the change in the severity of the patient’s neurological signs and symptoms from Video A to Video B. To investigate both the symptomatic and disease-modifying effects of treatment, N-acetyl-L-leucine is assessed during two treatment sequences: a 6-week parent study, and one-year extension phase.

**Discussion:** The novel CI-CS assessment, developed through a collaboration of all stakeholders, is advantageous in that it better ensures the primary endpoint is functionally relevant for each patient, is able to capture small but meaningful clinical changes critical to the patients’ quality of life (fine-motor skills; gait), and blinds the primary outcome assessment. The results of these three trials will inform whether N-acetyl-L-leucine is an effective treatment for NPC, GM2, and A-T, and can also serve as a new therapeutic paradigm for the development of future treatments for other orphan diseases.

**Trial registrations:** The three trials (IB1001-201 for Niemann-Pick disease type C (NPC); IB1001-202 for GM2 Gangliosidoses (Tay-Sachs and Sandhoff); IB1001-203 for Ataxia-Telangiectasia (A-T)) have been registered at www.clinicaltrials.gov (NCT03759639; NCT03759665; NCT03759678), www.clinicaltrialsregister.eu (EudraCT: 2018-004331-71; 2018-004406-25; 2018-004407-39) and https://www.germanctr.de (DR KS-ID: DRKS00016567; DRKS00017539; DRKS00020511).

## Background

The Orphan Drug Act of 1983 [1] defines rare (“orphan”) diseases as those which affect 200,000 or fewer individuals in the United States. Recently, advances in diagnostic techniques, particularly next generation sequencing of DNA, have led to the rapid expansion in the number of recognized “ultra-rare diseases”, proposed to describe disorders with a prevalence of less than 1:100,000 individuals [2]. It is now estimated there are over 10,000 rare and ultra-rare diseases which, although individually rare in prevalence, are collectively estimated to affect some 30 million Americans [3]. For over 95% of these disorders, there are no US Food and Drug Administration (FDA) approved treatments [4].

The lack of approved orphan-drugs reflects the unique challenges of orphan drug development. The conventional pathway to drug approval is often unsuitable for rare diseases, and raises hurdles that cannot be easily overcome, if at all. [5,6]. Novel drug-development strategies are needed to render the regulatory and development processes more feasible and cost-effective, and more quickly address the extremely-high unmet medical need of orphan diseases [7].

One response to expedite the development of novel treatments for rare diseases is to utilize a master protocol, where a single drug can be investigated for multiple indications. These may take three forms:

i. Umbrella trials, which study multiple targeted therapies in a single disease;
ii. Platform trials, in which multiple targeted therapies are studied in a single disease on an ongoing basis. Therapies are added or subtracted to the platform on the basis of previously agreed algorithms, with therapies allowed to enter or leave the platform on the basis of a decision algorithm;
iii. Basket studies, in which a single targeted therapy is studied in multiple diseases, or disease subtypes [8].

Here, we describe the development of a master protocol comprising three separate basket studies to investigate a novel agent, N-acetyl-L-leucine (Sponsor Code: IB1001), as a therapy for three different ultra-rare neurodegenerative diseases: Niemann-Pick disease, type C (NPC), GM2 Gangliosidoses (Tay-Sachs and Sandhoff diseases; “GM2”), and Ataxia-Telangiectasia (A-T), which share cerebellar ataxia as a common central manifestation.

### Protocol references

**IB1001-201** investigates N-acetyl-L-leucine for the treatment of Niemann-Pick disease type C (NPC). NPC is an ultra-rare (1:120,000), prematurely fatal, autosomal recessive, neurovisceral lysosomal storage disease that predominantly affects children. However, adolescent and adult onset cases are being increasingly recognized [9,10]. Treatment of NPC is so far limited to reducing the rate of disease progression with the substrate reduction therapy drug miglustat (Zavesca™ [11]), approved in the European Union and several other countries, but not in the United States.

**IB1001-202** investigates N-acetyl-L-leucine for the treatment of the GM2 Gangliosidoses (Tay-Sachs and Sandhoff diseases; “GM2”). GM2 is an ultra-rare (0.28:100,000), prematurely-fatal, autosomal recessive, neurovisceral lysosomal storage disorder that predominantly and most severely affect pediatric patients [12]. There is currently no approved treatment in any jurisdiction for GM2.

**IB1001-203** investigates N-acetyl-L-leucine for the treatment of Ataxia Telangiectasia (A-T). A-T is an ultra-rare (1:40,000-100,000), autosomal recessive, cerebellar ataxia that predominantly affects pediatric patients [13]. There is currently no approved treatment in any jurisdiction for A-T.

Despite their different aetiologies, NPC, GM2, and A-T are all characterized by progressive neurodegeneration of the cerebellum and cerebrum, resulting in physical and cognitive decline, and premature death. Each disorder manifests systemic, neurological, and neuropsychological findings. The three disorders share a number of hallmark symptoms, in particular, cerebellar ataxia, dysarthria, and dysphagia. Owing to their common neurological manifestations, and the mechanism of action of the investigational product N-acetyl-L-leucine, a single master protocol has been developed for the three disorders.

### N-acetyl-L-leucine

N-acetyl-L-leucine is the L-enantiomer of N-acetyl-leucine, a modified amino acid that has been available in France since 1957 as a racemate (equal amounts of both D- and L-enantiomers) under the trade name Tanganil^®^ (Pierre Fabre Laboratories) as a treatment for acute vertigo. N-acetyl-L-leucine is not approved for any indication in any jurisdiction.

The mechanisms of action of N-acetyl-DL-leucine for vertigo are not fully understood. It may act directly on neurons; in the vestibular nuclei, N-acetyl-DL-leucine has been shown to restore membrane potential in hyperpolarized/depolarized vestibular neurons following unilateral labyrinthectomy in guinea pigs [14]. This effect may be mediated by N-acetyl-DL-leucine’s direct interactions with membrane phospholipids such as phosphatidylinositol 4,5-bisphosphate, which influence ion channel activity [15]. Thus, acetyl-DL-leucine can normalize neuronal function. In patients with a unilateral neurotomy and labyrinthectomy, the agent was described to normalize the vestibular asymmetry, showing an effect only in the subgroup of individuals with residual vestibular function [16].

Subsequent studies in models of vertigo on the individual enantiomers have revealed that the therapeutic effects of N-acetyl-DL-leucine are due to the L-enantiomer. Studies of a rat model of unilateral labyrinthectomy revealed that N-acetyl-L-leucine, but not N-acetyl-D-leucine, is the pharmacologically active substance that improves central vestibular compensation [17]. Furthermore, a study in a unilateral vestibular neurectomy cat model suggested that N-acetyl-L-leucine is the enantiomer that leads to a significant acceleration of the vestibular compensation process, most likely acting on vestibular nuclei neurons [18].

Given the phylogenetic and electrophysiological similarities and close interactions between vestibular and deep cerebellar neurons, it was hypothesized both N-acetyl-DL-leucine and therefore also N-acetyl-L-leucine may have clinical utility in the treatment of cerebellar symptoms that occur in diseases such as NPC through a similar mechanism as that observed in models of vertigo, acting on neurons. However, i*n vitro* experiments using non-neuronal, *Npc1*-deficient Chinese Hamster Ovary cells or fibroblasts derived from patients with NPC, demonstrated N-acetyl-L-leucine and N-acetyl-DL-leucine also reverse disease-related cellular phenotypes in non-neuronal cells, including expanded lysosomal volume, with superior efficacy resulting from the L-enantiomer [19]. The mechanisms leading to effects on lysosomal storage in non-neuronal cells are currently under investigation.

N-acetyl-L-leucine has also been demonstrated to reduce neuroinflammation in the cerebellum. Activated microglia are associated with the neurodegenerative and neuroinflammatory components of cerebellar disorders, as engulfment of neuronal processes by microglia precede Purkinje cell death (an important type of nerve cell involved in movement control). *In vivo* studies in a mouse model for Traumatic Brain Injury demonstrate treatment with N-acetyl-L-leucine improves lysosome-related autophagy flux, and thereby restores its neuroprotective function in the cortices after traumatic brain injury [20]. This is expected to lead to the attenuation and restrict neuronal cell death, hence improving neurological function [20]. As acute and prolonged neuroinflammation contribute to neuronal death, these results suggest that N-acetyl-L-leucine may protect cells from neurodegeneration arising from genetic or acquired neurological disorders.

### Trial Rationale

The development of N-acetyl-leucine for NPC, GM2, and A-T began with the investigation of the commercially available racemic mixture, N-acetyl-DL-leucine (Tanganil^®^). In a case series of 12 patients with NPC, it was shown that N-acetyl-DL-leucine (3 g/day for one week followed by 5 g/day for three weeks) significantly improved the symptoms of NPC after 4-weeks of treatment, measured by the Scale for the Assessment and Rating of Ataxia (SARA), the Spinocerebellar Ataxia Functional Index (SCAFI) and EuroQol-5D-5L. N-acetyl-DL-leucine was well tolerated, and no side effects except for intermittent dizziness, were reported [21]. An additional case series describes the disease-modifying effect of long-term treatment with N-acetyl-DL-leucine in 10 patients with NPC treated for a median length of 7.7 months (maximum 21.2, minimum 2.7 months) [22].

The clinical utility of N-acetyl-DL-leucine for the treatment of Tay-Sachs, Sandhoff, and A-T has also been investigated in a compassionate-use case-series, as well as a variety of inherited cerebellar ataxias [23–25].In all studies, the compound was well-tolerated with no discernible serious side effects.

These preliminary clinical findings have been supported by *in vitro* studies in NPC and GM2 Gangliosidoses patient cell lines, and *in vivo* studies in the NPC (*Npc1^-/-^*) and Sandhoff (*Hexb*^*-/-*^) mouse models, which corroborate the pharmacological properties of N-acetyl-L-leucine and N-acetyl-DL-leucine in relation to the observed therapeutic effects (Ecem Kaya and Frances Platt, personal communication) [19].

The L-enantiomer, i.e. acetyl-L-Leucine, is believed to have potential clinical benefits compared to the racemic mixture [14,15, 16]. Further, pharmacokinetic studies suggest that the D-enantiomer could accumulate relative to the L-enantiomer during chronic administration of the racemate, which has the potential for long term negative effects [26].

### Rare disease trial design

The development of N-acetyl-leucine has been prioritized for the treatment of NPC, GM2, and A-T based on the existing pre-clinical and clinical data and the high unmet medical need of each indication. NPC, GM2, and A-T are progressive, life-threatening conditions with limited or no approved drugs, mandating greater urgency for trials to be conducted as efficiently as possible to maximize the chance they can be made available before the window of therapeutic opportunity is lost. However, like many clinical trials for rare and ultra-rare diseases, there are a number of critical protocol considerations which must be considered to do so:

1. For each disorder, the potential pool of participants is small, and within that circumscribed group, not all individuals are willing to participate or suitable candidates for clinical trials.
2. Parents and caregivers in these communities have legitimate ethical concerns about placebo-controlled trials. This complicates and delays the recruitment and completion of studies, and subsequently, approval and availability of the treatment for patients. The risk is even greater for trials like IB1001 which involves a modified version of a compound (Tanganil^®^) that is approved in France (as well as Lebanon, Vietnam, and Tunisia) for use in another clinical setting (acute vertigo), and which is known to be widely used in an unlicensed setting within these patient communities.
3. Each of these indications is characterized by broad variability in the symptoms and signs of rare diseases they exhibit, in the age at which they first present, and the rate at which they progress. The high inter-individual variability in the clinical course of the disease renders an assembly of well-matched cohorts of patients for controlled trials impossible.
4. Given the heterogeneity of the diseases, there are significant limitations in selecting and prioritizing a single outcome measure that can be considered clinically meaningful for the entire patient population.

Together, these factors significantly diminish a study’s ability to detect a therapeutic effect. Thus, detecting a statistically significant difference between intervention and control groups is hard to achieve.

### Collaboration

As a consequence of these challenges, many promising treatments for orphan diseases will never surpass the development hurdles and become approved for patients. In too many instances, when a rare disease trial fails, it is not clear if this is a consequence of the compound’s lack of a biological effect, or an inadequate study design that was not compatible with what can be reasonably achieved within the rare disease patient population. To avoid this pitfall in the IB1001 studies, an innovative master protocol and novel primary outcome measure were developed through a close collaboration between Regulatory Agencies, Key Opinion Leaders – including clinicians and patient communities—and the industry sponsors. Such cooperation was essential to create a trial design founded on a strong scientific rationale, but also taking into account the demographics of these heterogenous, rare populations. It is our hope that the collaboration between these stakeholders, and resulting development program for IB1001, represent a model to expedite the development of novel treatments for rare diseases.

## Methods/design

### Trial centers

The IB1001-201 (NPC), IB1001-202 (GM2) and IB1001-203 (A-T) clinical trials are separate multinational, multicenter, open-label, rater-blinded phase II studies. Each trial will enroll approximately 30 patients. Subjects are currently screened at 13 centers across Germany, Spain, Slovakia, the United Kingdom and the United States, including Ludwig Maximilians University, Munich (201, 202, 203); University of Giessen (201, 202, 203); Bellvitge University Hospital (201, 202); Comenius University in Bratislava (201); Birmingham Women’s and Choldren’s NHS Foundation Trust (203); Great Ormond Street Hospital (201); Royal Free Hospital (201); Royal Manchester Children’s Hospital (201, 202); Salford Royal NHS Foundation Trust (201, 202); Royal Papworth Hospital (203); The Mayo Clinic, Rochester MN (201, 202); New York University Langone (202) and University of California Los Angeles (202, 203).

### Study oversight

The IB1001 trials are conducted in accordance with the International Conference for Harmonisation (of Technical Requirements for Pharmaceuticals for Human Use) - Good Clinical Practice Guideline, the General Data Protection Regulator, and the Declaration of Helsinki. The studies have been approved by the ethics committees of each participating center and the regulatory authorities in each respective country. The safety, integrity, and feasibility of the trial is monitored by an independent data safety monitoring board (DSMB) consisting of three independent, non-participating members (including two clinicians and a statistician). The function of the DSMB is to monitor the course of the studies and, as applicable, give a recommendation to the Sponsor of the trial for discontinuation, modification or continuation of the study.

### Patient population and eligibility criteria

Patients are screened for eligibility according to the inclusion and exclusion criteria. To be eligible for the respective study, patients with a confirmed diagnosis of NPC, GM2, or A-T (aged ≥6 years in Europe and ≥18 years in the US) must present with clinical symptoms, provide appropriate informed consent, and undertake a washout of any prohibited medications (if applicable). These include any variant of N-acetyl-DL-leucine (e.g. Tanganil^®^). For a detailed description of the inclusion and exclusion criteria see **Table 1**.

**Table 1.** Inclusion and exclusion criteria for patient selection in the Parent Study.

| Inclusion Criteria | Exclusion Criteria |
| --- | --- |
| Individuals who meet all of the following criteria are eligible to participate in the study:<br>Written informed consent signed by the patient and/or their legal representative/ parent/ impartial witness<br>**IB1001-201 (NPC) EU:** Male or female aged $\geq 6$ years in Europe OR $\geq 18$ years in the United States with a confirmed diagnosis of NPC at the time of signing informed consent. Confirmed diagnosis includes<br>Clinical features and positive biomarker screen and/or filipin test without genetic tests results (has not been performed)<br>Clinical features and positive genetic test<br>Clinical features and positive biomarker screen and/or filipin test but only one NPC mutation identified on genetic test<br>Clinical features with positive biomarker screen and/or filipin test and positive genetic test<br>**IB1001-201 (NPC) US:** Male or female aged $\geq 6$ years in Europe OR $\geq 18$ years in the United States with a confirmed diagnosis of NPC at the time of signing informed consent. Patients must have clinical features of NPC and a positive genetic test for mutations in both copies of NPC1 or in both copies of NPC2.<br>**IB1001-202 (GM2):** Male or female aged $\geq 6$ years in Europe OR $\geq 18$ years in the United States with a confirmed diagnosis of GM2 Gangliosidosis at the time of signing informed consent. Confirmed diagnosis, i.e., clinical features and positive genetic test GM2-gangliosidosis caused by β-hexosaminidase deficiency resulting from mutations in the HEXA or HEXB genes<br>**IB1001-203 (A-T):** Male or female aged $\geq 6$ years in Europe OR $\geq 18$ years in the United States with a confirmed genetic diagnosis of A-T at the time of signing informed consent. | Individuals who meet any of the following criteria are not eligible to participate in the study:<br>Asymptomatic patients<br>Patient has clinical features of NPC and a positive biomarker screen and/or filipin test, but a completely negative result on a previous genetic test for NPC<br>Patients who have any of the following:<br>Chronic diarrhea;<br>Unexplained visual loss;<br>Malignancies;<br>Insulin-dependent diabetes mellitus.<br>Known history of hypersensitivity to the Acetyl-Leucine (DL-, L-, D-) or derivatives.<br>History of known hypersensitivity to excipients of Ora-Blend® (namely sucrose, sorbitol, cellulose, carboxymethylcellulose, xanthan gum, carrageenan, dimethicone, methylparaben, and potassium sorbate).<br>Simultaneous participation in another clinical study or participation in any clinical study involving administration of an investigational medicinal product (IMP; ‘study drug’) within 6 weeks prior to **Visit 1**.<br>Patients with a physical or psychiatric condition which, at the investigator’s discretion, may put the patient at risk, may confound the study results, or may interfere with the patient’s participation in the clinical study.<br>Known clinically significant (at the discretion of the investigator) laboratories in hematology, coagulation, clinical chemistry, or urinalysis, including, but not limited to:<br>**IB1001-201/ IB1001-202:** Aspartate aminotransferase (AST) or alanine aminotransferase (ALT) $>5x$ upper limit of normal (ULN);<br>**IB1001-203:** Aspartate aminotransferase (AST) or alanine aminotransferase (ALT) $>3x$ upper limit of normal (ULN);<br>Total bilirubin $>1.5x$ ULN, unless Gilbert’s syndrome is present in which case total bilirubin $>2x$ ULN.<br>Known or persistent use, misuse, or dependency of medication, drugs, or alcohol. |
| 3. Females of childbearing potential, defined as a premenopausal female capable of becoming pregnant, will be included if they are either sexually inactive (sexually abstinent<sup>1</sup> for 14 days prior to the first dose and confirm to continue through 28 days after the last dose) or using one of the following highly effective contraceptives (i.e. results in <1% failure rate when used consistently and correctly) 14 days prior to the first dose continuing through 28 days after the last dose:<br>a) intrauterine device (IUD);<br>b) surgical sterilization of the partner (vasectomy for 6 months minimum);<br>c) combined (estrogen or progestogen containing) hormonal contraception associated with the inhibition of ovulation (either oral, intravaginal, or transdermal);<br>d) progestogen only hormonal contraception associated with the inhibition of ovulation (either oral, injectable, or implantable);<br>e) intrauterine hormone releasing system (IUS);<br>f) bilateral tubal occlusion.<br>4. Females of non-childbearing potential must have undergone one of the following sterilization procedures at least 6 months prior to the first dose:<br>a) hysteroscopic sterilization;<br>b) bilateral tubal ligation or bilateral salpingectomy;<br>c) hysterectomy;<br>d) bilateral oophorectomy;<br>**OR** be postmenopausal with amenorrhea for at least 1 year prior to the first dose and follicle stimulating hormone (FSH) serum levels consistent with postmenopausal status. FSH analysis for postmenopausal women will be done at screening. FSH levels should be in the postmenopausal range as determined by the central laboratory.<br>5. Non-vasectomized male patient agrees to use a condom with spermicide or abstain from sexual intercourse during the study until 90 days beyond the last dose of study medication and the female partner agrees to comply with inclusion criteria 3 or 4. For a vasectomized male who has had his vasectomy 6 months or more prior to study start, it is required that they use a condom during sexual intercourse. A male | 8. Current or planned pregnancy or women who are breastfeeding.<br>9. Patients with severe vision or hearing impairment (that is not corrected by glasses or hearing aids) that, at the investigator's discretion, interferes with their ability to perform study assessments.<br>10. Patients who have been diagnosed with arthritis or other musculoskeletal disorders affecting joints, muscles, ligaments, and/or nerves that by themselves affects patient's mobility and, at the investigator's discretion, interferes with their ability to perform study assessments.<br>11. Patients unwilling and/or not able to undergo a 42-day washout period from any of the following prohibited medication prior to **Visit 1** (Baseline 1) and remain without prohibited medication through **Visit 6**.<br>a) Aminopyridines (including sustained-release form);<br>b) N-Acetyl-DL-Leucine (e.g. Tanganil<sup>®</sup>) ;<br>c) N-Acetyl-L-Leucine (prohibited if not provided as IMP);<br>d) Riluzole;<br>e) Gabapentin;<br>f) Varenicline;<br>g) Chlorzoxazone;<br>h) Sulfasalazine;<br>i) Rosuvastatin. |
| who has been vasectomized less than 6 months prior to study start must follow the same restrictions as a non-vasectomized male.<br>6. If male, patient agrees not to donate sperm from the first dose until 90 days after their last dose.<br>7. Patients must fall within:<br>a) A SARA score of $5 \leq X \leq 33$ points (out of 40)<br>**AND**<br>b) Either:<br>i. Within the 2-7 range (0-8 range) of the Gait subtest of the SARA scale<br>**OR**<br>ii. Be able to perform the 9-Hole Peg Test with Dominant Hand (9HPT-D) (SCAFI subtest) in $20 \leq X \leq 150$ seconds.<br>8. Weight $\geq 15$ kg at screening.<br>9. Patients are willing to disclose their existing medications/therapies for (the symptoms) of NPC, including those on the prohibited medication list. Non-prohibited medications/therapies (e.g. miglustat, concomitant speech therapy, and physiotherapy) are permitted provided:<br>a) The Investigator does not believe the medication/therapy will interfere with the study protocol/results<br>b) Patients have been on a stable dose/duration and type of therapy for at least 42 days before **Visit 1** (Baseline 1)<br>c) Patients are willing to maintain a stable dose/do not change their therapy throughout the duration of the study.<br>10. An understanding of the implications of study participation, provided in the written patient information and informed consent by patients or their legal representative/parent, and demonstrates a willingness to comply with instructions and attend required study visits (for children this criterion will also be assessed in parents or appointed guardians). |  |

<sup>1</sup> Sexual abstinence is considered a highly effective method only if defined as refraining from heterosexual intercourse during the entire period of risk associated with the study treatments. In this trial abstinence is only acceptable if in line with the patient's preferred and usual lifestyle.
Periodic abstinence (calendar, symptothermal, post-ovulation methods), withdrawal (coitus interruptus), spermicides only, and lactational amenorrhoea method (LAM) are not acceptable methods of contraception. As well, female condom and male condom should not be used together.

### Recruitment and patient involvement

Patients are recruited via personal correspondence, routine care appointments, and referrals. In addition, there is tremendous collaboration and support from multinational patient organizations representing these rare disease communities.

All eligible patients who agree to participate in the study are provided with a full verbal explanation of the trial and the Patient Information Sheet. This includes detailed information about the rationale, design, and personal implications of the study.

### Study design and procedures

The IB1001 clinical trials are open-label, rater-blinded studies. During the development of the IB1001 master protocol, the appropriateness of initiating a randomized, double-blind, controlled studies for these ultra-orphans was strongly questioned by clinical experts and representatives of the patient communities, given the diseases’ relentless and often rapid progression, prematurely fatal outcome, and lack of alternative treatment options. A formal feasibility study was initiated by the Sponsor that demonstrated that a randomized, double-blind, placebo-controlled, crossover study would be difficult to recruit and carry out without changes to the study design. This was largely attributed to the known, widespread, off-label/unlicensed use of N-acetyl-DL-leucine (Tanganil^®^). Patients and families expressed reluctance to participate in a placebo-controlled study where they would be required to washout from this off-label/unlicensed medication and receive an inactive treatment for even 50% of the time.

In order to assure the feasibility of recruitment, an open-label study schema is used. Based on observational and pre-clinical studies that demonstrate the potential of symptomatic benefit of treatment in as little as 4-weeks [21,23], in the first treatment sequence (“Parent Study”), patients are assessed during three study phases: a baseline period (with or without a study run-in), a treatment period of 6-weeks (42-49 days), and a washout period of 6-weeks (42-49 days). Patients will be assessed twice during each period to allow an assessment of intra-patient variability.

At the initial screening visit, patients will be classified as either “naïve” or “non-naïve” depending on their use of prohibited medications within the past 6-weeks (42 days). The schedule of events during the initial screening visit and throughout the baseline period (through Visit 1) will vary depending on the patient’s classification as either “naïve” or “non-naïve”.

Given the known unlicensed use of the racemate (Tanganil^®^), for all patients, a urine sample will be taken at Visit 1 to detect N-acetyl-D-leucine using a validated Liquid Chromatography Mass Spectrometry/Mass Spectrometry method. Provided the level of N-acetyl-D-leucine tests below the permitted threshold, the initial screening visit will be confirmed as Visit 1 (Baseline 1). If a patient classified as “Naïve” unexpectedly tests positive for levels of N-acetyl-D-leucine above the permitted threshold, at the direction of their Principal Investigator, a run-in wash-out period of 6-weeks (42 days) is requested before they are eligible to return for a repeat Visit 1. Patients who fail two urine N-acetyl-D-leucine tests (e.g. Visit 1 and repeat Visit 1) are ineligible for the study.

**Figure 1** displays the naïve and non-naïve study schemes for the Parent Study. **Suppl. Table 1** lists the schedule of enrolment and assessments together with pre-planned time points for clinic visits during Patients who have completed Visit 6 of the Parent Study have the opportunity to continue treatment with N-acetyl-L-leucine (IB1001) in an Extension Phase if the Principal Investigator determines it is in their best interest. The Extension Phase consists of a one-year (351-379 day) treatment period followed by a 6-weeks (42-56 days) washout period. **Table 2** lists the inclusion criteria for the Extension Phase. **Figure 2** displays the Extension Phase study schema. **Suppl. Table 2** lists the schedule of enrolment and assessments together with pre-planned time points for clinic visits in the Extension Phase.

**Table 2.** Inclusion criteria for patient participation in the Extension Study.

| Inclusion Criteria |
| --- |
| 1. Completed Visit 6 of the IB1001-201 / IB1001-202 / IB1001-203 Parent Study<br>2. The Principal Investigator determines further treatment with IB1001 to be in patient's best interest<br>3. Written informed consent signed by the patient and/or their legal representative/parent/ impartial witness for participation in the Extension Phase<br>4. Patients are willing to continue to remain without the following prohibited medication from **Visit 6** throughout the duration the Extension Phase:<br>j) Aminopyridines (including sustained-release form);<br>k) N-Acetyl-DL-Leucine (e.g. Tanganil®);<br>l) N-Acetyl-L-Leucine (prohibited if not provided as investigational medicinal product [IMP]);<br>m) Riluzole;<br>n) Gabapentin;<br>o) Varenicline;<br>p) Chlorzoxazone;<br>q) Sulfasalazine;<br>r) Rosuvastatin. |

**Figure 1.**
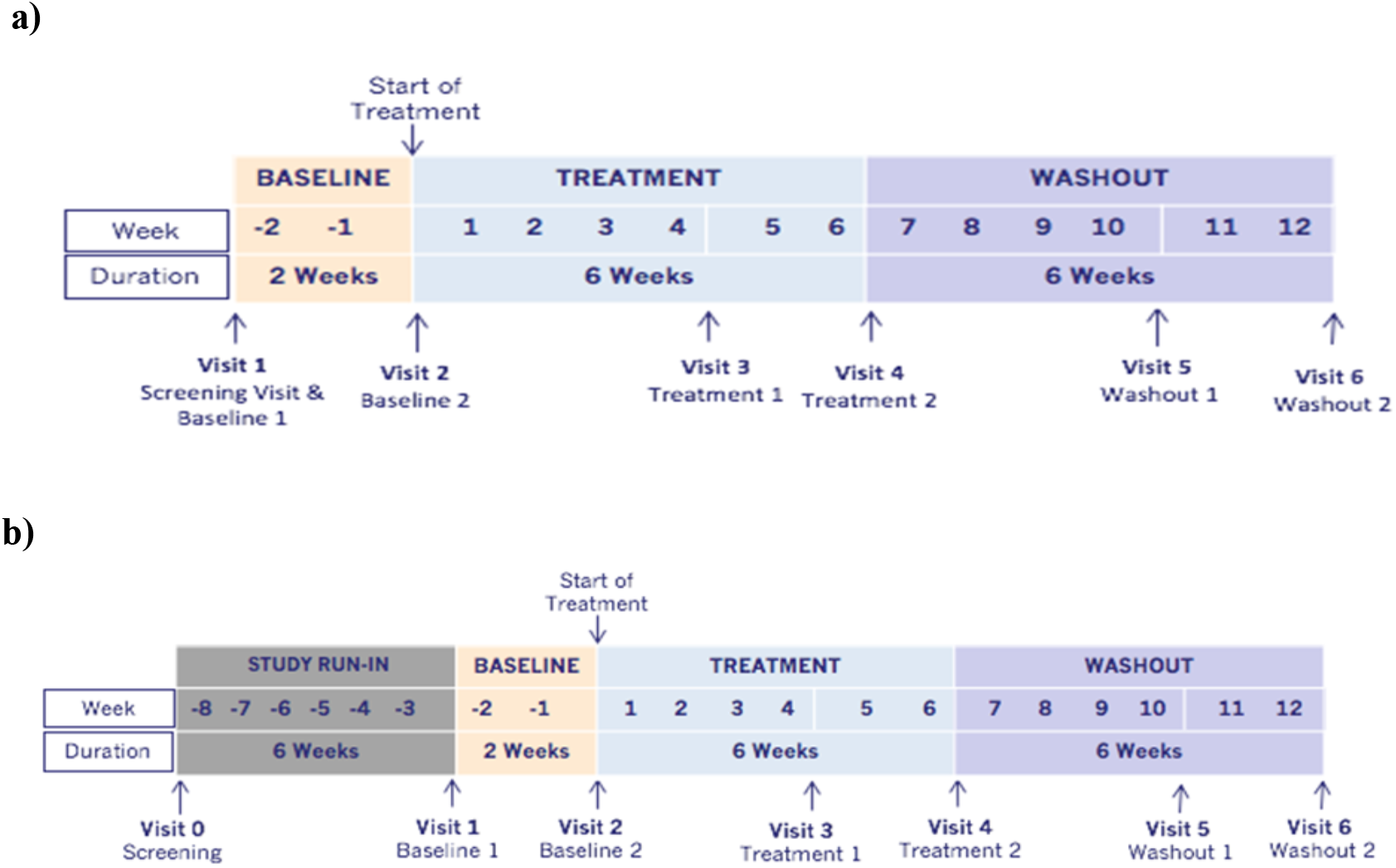
Parent Study Schema. During the treatment period, all patients will receive N-Acetyl-L-Leucine for 42 days (+7 days). Visit 3 (Treatment 1) will occur at Day 28 (+7 days) of the treatment period and Visit 4 (Treatment 2) will occur after the full 42 days (+7 days) of treatment. A 42-day (+7 days) washout period will be performed following treatment with N-Acetyl-L-Leucine. Visit 5 (Washout 1) will occur on Day 28 (+7 days) of the washout period and Visit 6 (Washout 2) will occur after the full 42 days (+7 days) of washout. **a)** Naïve patients screening pathway: patients who have not used any prohibited medications within 42 days of screening are “naïve”. Their initial screening visit is treated as Visit 1 (Baseline 1). **b)** Non-naïve patient screening pathway: patients who have used or are unable to confirm or deny if they have used, any prohibited medication within the past 42 days are “non-naïve”. Patient will be given the opportunity to undergo a minimum of 42 days washout before returning for a repeat Visit 1 (Baseline 1).

**Figure 2.**
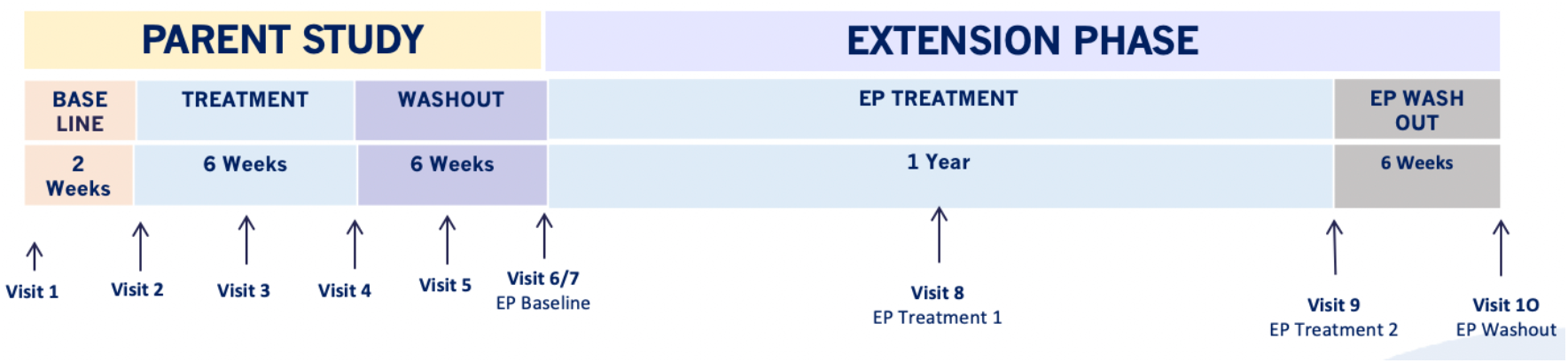
Extension Phase Schema. Patients will be assessed approximately 4 times over a 64-week period: at the start of the Extension Phase, after 6 months of treatment, 1 year of treatment, and after a 42-day (+14 day) post-extension-phase treatment washout.

### Study drug

In the Parent Study, the dosage formulation of N-acetyl-L-leucine is 1,000 mg powder for oral suspension (manufacture: Patheon UK Limited, Oxfordshire United Kingdom) which is suspended in 40 mL Ora-Blend^®^.

In the Extension Phase, the dosage form of N-acetyl-L-leucine is 1,000 mg granules for oral suspension in a sachet (manufacture: Patheon UK Limited, Oxfordshire United Kingdom and Patheon France S.A.S., Bourgoin France) which is suspended in 40 mL water.

### Administration and study drug dosage

During the treatment periods for both treatment sequences, the dosing of the study drug is as follows: Patients aged ≥13 years or aged 6-12 years weighing ≥35 kg will take 4 g/day (2 g in the morning, 1 g in the afternoon, and 1 g in the evening). Patients aged 6-12 years weighing 25 to <35 kg will take 3 g /day (1 g in the morning, 1 g in the afternoon, and 1 g in the evening). Patients aged 6-12 years weighing 15 to <25 kg will take 2 g /day (1 g in the morning and 1 g in the evening). Medication should be taken at least 30 minutes before or at least 2 hours after a meal.

If adverse events are noted, patients are permitted to down-titrate to one-half their daily dose, at the direction of the investigator. Compliance will be assessed upon a review of the inventory of IB1001 bottles/sachets returned by patients.

### Study objectives

The two treatment sequences in the Parent Study and Extension Phase enable the investigation of both the symptomatic (6-week), and long-term (one-year) safety and efficacy of treatment with N-acetyl-L-leucine.

The primary objective of both treatment sequences is to evaluate the efficacy of N-acetyl-L-leucine based on blinded raters’ Clinical Impression of Change in Severity © (CI-CS) in the treatment of NPC, GM2, or A-T.

The secondary objectives are:

- To assess the clinical efficacy (symptomatic and long-term) of N-acetyl-L-Leucine on symptoms of ataxia, functioning, and quality of life for patients with NPC, GM2, or A-T;
- To evaluate the safety and tolerability of N-acetyl-L-leucine at 4 g/day in patients with NPC, GM2, or A-T, including patients aged ≥18 years in the United States and patients aged ≥13 years in Europe, and weight-tiered doses in patients 6 to 12 years of age in Europe.

In the Extension Phase, an additional secondary objective is to characterize the pharmacokinetics of N-acetyl-L-leucine in patients with NPC, GM2, or A-T.

### Safety and efficacy parameters

#### Primary efficacy endpoint

In light of the defined challenges to conducting an open-label clinical trial in these ultra-orphan diseases, a novel primary endpoint, the Clinical Impression of Change in Severity © (CI-CS) was developed.

The administration and assessment of the CI-CS involves three tasks. **Table 3** provides an overview of each task, the responsible party and the timepoint at which it is performed.

**Table 3.** Components of the administration and scoring of the Clinical Impression of Change in Severity^©^ (CI-CS) Assessment

| Task | Timepoint | Responsible Person(s) |
| --- | --- | --- |
| Determine CI-CS Primary Anchor Test (8MWT or 9HPT-D) | Visit 1 | Principal Investigator |
| Anchor Tests Video Recordings | Visit 2, 3, 4, 5, 6, 7, 8, 9, 10, Early Termination (as applicable) | Qualified Video-Recorder (Study Team Member) |
| CI-S & CI-CS Assessment Anchor Tests | Following Last Visit (Parent Study: Visit 6; Extension Phase: Visit 10) | Blinded Independent Raters |

##### (1) Determine CI-CS primary anchor test (8MWT or 9HPT-D)

Given the heterogeneity of symptoms, the appointment of a single symptom scale, such as either the 8 Meter Walk Test (8MWT) or the 9 Hole Peg Test of the Dominant Hand (9HPT-D), as the primary outcome measure is not appropriate for patient populations. To better ensure the primary outcome measure is clinically meaningful for each individual patient, at Visit 1, the Principal Investigator selects either the 8MWT or 9HPT-D to be the patient’s “Primary Anchor Test”. The selection is guided by pre-defined criteria based on patient’s performance on ataxia rating scales (Spinocerebellar Ataxia Functional Index (SCAFI) and Scale for the Assessment and Rating of Ataxia (SARA)), as well as interactions with the patient, their family, and/or caregivers. A cognitive assessment is also performed at Visit 1 according to the standard procedures of the clinical site to select an anchoring functional test which is appropriate for each patient from both a cognitive and a motor perspective. The primary anchor test remains fixed for each patient for the duration of the study.

##### (2) Video recordings: primary and non-primary anchor tests

At each study visit (except Visit 0), the 8MWT and 9HPT-D are videoed in a standardized manner by qualified members of the study team. Video consultation has been validated in other diseases and can minimize variability in the data [27–29]. Out of necessity, the trials are conducted at multiple study sites in different geographical locations, with inherent potential for considerable inter-rater variability in assessment scores [27]. Video recordings allow for centralized review and repeated viewing without the necessity for repetition and patient fatigue [30].

Each clinical site must qualify two videographers before any patients are screened. All patient videos should be performed by the same videographer, in the same environment, to ensure consistency and reduce the potential for variability. Videos will be collected by the clinical sites and submitted to Medpace Core Laboratories via the web based ClinTrak Imaging Windows Client Application for collection, quality control, central review, and storage. A more detailed description on the acquisition, submission, and video review process is provided in **Suppl. Material I**.

##### (3) CI-CS assessment of primary anchor test

The primary CI-CS assessment is performed by two, independent raters based on the videos of each patient’s primary anchor test acquired throughout the respective treatment sequence (Visit 1 – 6; Visit 7-10). Prior to their review of patient videos, the raters are trained on existing videos to ensure standardization of the assessments.

Raters are asked to assess three video pairs:

- Pair A: (i) Baseline and (ii) End of Treatment
- Pair B: (ii) End of Treatment and (iii) End of Washout
- Pair C: (iii) End of Washout and (i) Baseline

Pairs A, B, and C are reviewed by the raters in random order. The internal pairing of videos (i) Baseline, (ii) End of Treatment, and (iii) End of Washout, will also be arranged randomly. The raters will be blinded in this way to reduce detection and performance biases.

For each pair, raters are asked to assess: “Compared to the first video, how has the severity of the patient’s performance on the 8MWT or 9HPT-D changed (improved or worsened) as observed in the second video?” The CI-CS assessment is based on a 7-point Likert scale ranging from + 3 (significantly improved) to −3 (significantly worse). If there is a difference of one (1) point in the two primary blinded raters’ CI-CS scores for a specific video pairing, the two scores will be averaged. If there is a difference greater than one (1) point between the two primary blinded reviewers’ CI-CS scores for a specific video pairing, an adjudication rating will be triggered. In such cases, a third blinded rater will review the scores given from each of the two primary independent raters and determine which score is more accurate, that of rater A or rater B (adjudication by consensus). The adjudicator’s decision will provide the final score for that video assessment.

#### Primary endpoint definition

The primary endpoint is defined as:

##### Assessment A

the CI-CS comparing videos of the patient’s performance on the pre-defined anchor test at (i) the end of treatment versus (ii) baseline;

**minus**

##### Assessment B

the CI-CS comparing videos of the patient’s performance on the pre-defined anchor test at (i) the end of washout versus (ii) end of treatment.

The CI-CS achieves the following: First, detection and performance bias for the primary endpoint is reduced by the blinded, independent review. Second, each patient’s washout period (Assessment B) serves as a control arm to their treatment period (Assessment A). Third, videos increase reliability and minimize the burden on patients. Finally, the CI-CS is a platform to capture and assess small but clinically meaningful changes in patient’s performance, which relate to their level of functioning and quality of life, but which cannot be obtained from the quantitative timed 8MWT or 9HPT-D assessments. This is critical: although functioning typically improves with symptom reduction, these concepts are not always concordant [31]. For example, a change in gait velocity does not necessarily account for the way gait patterns deviate from normal (i.e. balance, variability, asymmetry, rhythm, posture, or, notably, the ability to walk unaided [23,30]), and therefore, a meaningful improvement in ambulation. Similarly, a faster 9HPT score does not necessarily capture a change in fine-motor skills, grip, or tremor wherein individuals may perform the test more carefully or precisely which results, paradoxically, in an increase in 9HPT time. Therefore, the CI-CS is a metric of clinical importance that cannot be obtained from traditional assessment measures.

In the Extension Phase, the primary endpoint is defined as success on the Clinical Impression of Change in Severity© (CI-CS) comparing videos of the primary anchor test at (i) the end of the Extension Phase treatment with N-acetyl-L-leucine (Visit 9) versus (ii) the Extension Phase baseline (Visit 7). Clinical benefit is defined as a CI-CS value of 0 (no change) or better (≥1, at least some improvement).

#### Secondary efficacy endpoints to supplement the analysis of the primary endpoint

Supportive secondary endpoints will be evaluated that directly supplement the analysis of the primary endpoint, including the independent raters’ Clinical Impression of Severity (CI-S). The raters will be given videos of the patient’s performance on the primary and non-primary anchor test acquired at each visit of the respective treatment sequence (Visits 1 to 6; Visit 7 to 10). This will enable the evaluation of both of the possible anchor tests and assess the appropriateness of the chosen primary anchor test with regard to its ability to function as a clinically meaningful outcome measure for the patient. Again, videos will be presented to the raters in a randomized, blinded manner.

For each video, the raters are asked to assess: *“Considering your total clinical experience with this particular population, how ill is the patient at this time?”* The CI-S is rated on a 7-point Likert scale ranging from + 3 (normal, not at all ill) to −3 (among the most extremely ill patients). For the CI-S, if there is any difference in the two blinded reviewers’ scores, the two scores will be averaged.

#### Secondary endpoints

Additional secondary endpoints will measure other symptoms and evaluate quality of life. Descriptive statistics will be provided for these measures at each visit, and also, changes from [Parent Study/Extension Phase]: baseline (Visit 2 / Visit 7) to the end of treatment with N-acetyl-L-leucine (Visit 4 / Visit 9), as well as end of treatment with N-acetyl-L-leucine (Visit 4 / Visit 9) to the end of the post-treatment washout (Visit 6 / Visit 10) for the following measures:

- Spinocerebellar Ataxia Functional Index (SCAFI) [32].
- Scale for Assessment and Rating of Ataxia (SARA) score [32,33].
- Quality of Life EQ-5D-5L for patients aged ≥18; EQ-5D-Y for patients aged <18 years [34].
- Modified Disability Rating Scale (mDRS) (201 and 202 study only) [35,36].
- Clinical Global Impression Scales [37]:
  ○ Investigator, Caregiver, and Patient (*if able)* Clinical Global Impression of Severity at every visit
  ○ Investigator, Caregiver, and Patient (*if able)* Clinical Global Impression of Improvement comparing end of treatment (Visit 4 / Visit 9) to baseline (Visit 2 / Visit 7), and end of washout (Visit 6 / Visit 10) to end of treatment (Visit 4 / Visit 9)
- Columbia Suicide Severity Rating Scale (203 study only) [38].

In the Extension Phase of the IB1001-201 study only, the Niemann-Pick disease type C Clinical Severity Scale (NPC-CSS) will be introduced as a secondary endpoint, where the graduation of changes is expected to detect clinically relevant changes in functioning / benefit after one year of treatment [39].

In addition, the Annual Severity Increment Score [22] will be assessed in the Extension Phase.

#### Safety parameters

Adverse events (serious and non-serious), concomitant drug and non-drug therapies, safety laboratory blood samples (hemoglobin, erythrocytes, hematocrit, thrombocytes, leukocytes, sodium, lactate dehydrogenase, potassium, creatinine, serum bilirubin level, aspartate aminotransferase, alanine aminotransferase, urea, alkaline phosphatase, follicle-stimulating hormone for postmenopausal women only) and urine samples (leukocytes, nitrite, urobilinogen, protein, pH, occult blood (erythrocytes, leucocytes), specific gravity, ketones, bilirubin, glucose) will be collected routinely throughout the study. Sparse pharmacokinetic blood sampling will be conducted at every visit (Visit 1-Visit 10). Blood samples for the quantification of N-acetyl-L-leucine in plasma will be obtained at Visit 7 and Visit 9. Urine samples will also be collected for concentrations of N-acetyl-D-leucine at the time points designated on the schedule of events (**Suppl. Table 1, 2**). At Visit 1, this urine sample serves as a key enrollment criterion testing for the use of the prohibited medication N-acetyl-DL-leucine. Vital signs, physical exams, height/weight, and electrocardiograms will also be collected at the time points designated on the schedule of events (**Suppl. Table 1, 2)**. A detailed description of the safety parameters is provided in **Suppl. Material II**.

#### Statistical planning and analyses

The Statistical Analysis Plan details the statistical methods for analysis for each of the three clinical trials.

The primary analysis population is the modified intention to treat (mITT) analysis set defined as all patients who receive at least one dose of study drug (N-acetyl-L-leucine) and with one video recording at either Visit 1 or Visit 2 (or both). Analyses will also be conducted on the per protocol analysis set which will consist of all patients with video recordings at baseline (Visit 1 and/or Visit 2), end of treatment (Visit 3 and/or Visit 4), and end of washout (Visit 5 and/or Visit 6) and without any major protocol deviations that can influence the validity of the data for the primary efficacy variable.

The primary endpoint will utilize assessments based on single video recordings at the end baseline period (Visit 2), end of the treatment period (Visit 4), and end of the washout period (Visit 6). If the Visit 2 video is missing, the Visit 1 video will be used in its place. Similarly, if the Visit 4 and Visit 6 videos are missing Visit 3 and Visit 5 videos will respectively be used in their place. Analyses based on the mITT analysis set will utilize a last observation carried forward approach for missing videos 3 and 4 and missing videos 5 and 6. For the primary endpoint CI-CS, this implies that the CI-CS value for Visit 2 to Visit 4 will be assigned the value 0 if both videos 3 and 4 are not available, similarly for Visit 4 to Visit 6 if either both videos 3 and 4 or both videos 5 and 6 are unavailable.

The analysis of the primary endpoint will be based on a single sample one-sided t-test comparing the mean of the CI-CS differences with zero. The null hypothesis is that the mean is ≤0, with the alternative hypothesis that this mean is >0 and the test will be conducted at the one-sided 5% significance level. The secondary endpoint that is based on a 3-point categorization of the CI-CS will be evaluated based on the Wilcoxon-signed-rank test. The analysis of the secondary endpoint, Clinical Impression of Severity (CI-S) between baseline (average for Visit 1 and Visit 2) and end of treatment with N-acetyl-L-leucine (average for Visit 3 and Visit 4) minus the change in CI-S between end of treatment with N-acetyl-L-leucine (average for Visit 3 and Visit 4) and end of washout (average for Visit 5 and Visit 6), will be analyzed as for the primary endpoint. All other endpoints will be evaluated descriptively. There will be no formal control of multiplicity across these endpoints.

For each of the primary and secondary endpoints, there will be evaluation within key subgroups; naïve versus non-naïve, age (pediatric versus adult), gender (male versus female), region (US versus Europe), primary anchor test (9HPT-D or 8MWT), patients on miglustat vs patients not on miglustat (201 study only); Tay-Sachs versus Sandhoff patients (202 study only), individual components of SARA scale at baseline: Gait Subtest, composite of SARA Subtests 1-4 (Gait, Stance, Sitting, Speech), intra-patient variability between SARA score at Visit 1 (Baseline 1) versus Visit 2 (Baseline 2) (below/above median) and intra-patient variability between CI-S score Visit 1 (Baseline 1) versus Visit 2 (Baseline 2) (below/above median). These evaluations will be based on plotting treatment differences together with 90% confidence intervals within each subgroup.

It is postulated that N-acetyl-L-leucine will show effectiveness in 30% of patients and this success rate is viewed as being clinically important. It is assumed that this group of patients will have scores that are distributed across the values 1 and 2 for the primary endpoint with 10% scoring 1 and 20% scoring 2, and further that the remaining 70% of patients will have scores that are evenly distributed between the values −1, 0, and 1. The resulting mean score is 0.5 and the standard deviation for the primary endpoint under these assumptions is then 1.075. With 30 patients reporting on the primary endpoint, the study will have 80% power to detect a treatment benefit in a 5% one-sided one-sample t-test. Assuming alternatively that the 30% of patients improving will have scores that are evenly distributed across the values 1 and 2, then the mean score for the primary endpoint will be 0.45 with an SD of 1.02. Under these assumptions, the study will have power of 76%.

In observational case-series, N-acetyl-leucine has demonstrated potential as a broad treatment for the general symptoms of other progressive, inherited ataxias [23,24]. Meta-analyses will therefore be conducted across the three separate studies for NPC, GM2, and A-T to assess more general question regarding the effectiveness of N-acetyl-L-leucine for symptoms of ataxia. A description of Data Collection is provided in **Suppl. Material III**.

## Discussion

Given the lack of global symptomatic or disease-modifying therapies for NPC, GM2, or A-T, there is an urgent need for effective and well-tolerated drug treatments. The open-label, rater-blinded IB1001 master protocol was designed through a collaboration between National Regulatory Agencies, leading Clinical Experts, Patient Organizations, and the industry Sponsor, to address of the unique ethical and practical challenges to conducting clinical trials for these orphan, heterogenous patient populations, and be better able to capture N-acetyl-L-leucine’s therapeutic effect, and therefore, best positioned to expedite the development and availability of this promising drug candidate [7].

In Sponsor meetings with National Regulatory Authorities, methodological and statistical concerns were raised and discussed, with an emphasis on selecting a specific primary outcome measure that focuses on the aspects of the diseases that are relevant and important to patients. Subsequent interviews with representatives of the patient communities and leading clinicians, regarding the core signs/symptoms that are most meaningful for patients and clinically relevant to physicians, elucidated the key symptoms that affect patient’s quality of life. In addition, the active support of patient organizations throughout the Sponsor’s formal feasibility study allowed centers of excellence best capable of recruitment to be quickly identified. However, it also revealed the formidable barrier that the off-label, unlicensed use of the racemate (Tanganil^®^) would be to recruitment in a double-blind, placebo-controlled trial.

Ultimately, the issues raised during regulatory review and the feasibility process were instructive to developing the innovative trial design and adaptive primary outcome assessment for the IB1001 studies (tailored to the capabilities of individual patients to maximize inclusion, and better detect a clinically meaningful treatment effect). The pathway to the studies approval was facilitated by frequent communication between all parties, and the collaborative adaptation of study methodology and statistical approaches ensured the IB1001 trials were feasible to recruit, tailored to the capabilities of the NPC, GM2, and A-T patents, and importantly, best positioned to detect a meaningful clinical change in patients’ quality of life.

### Trial status

At the time of manuscript submission, the protocol for IB1001-201, 202, 203 have been accepted / approved in each country where they are respectively planned to be conducted, including: the US Food and Drug Administrations (201, 202, 203); UK Medicines and Healthcare products Regulatory Authority (201, 202, 203); German Federal Institute for Drugs and Medical Devices (201, 202, 203) Spanish Agency of Medicines and Medical Products (201, 202), Slovakia Štátny ústav pre kontrolu liečiv (201), as well as independent ethics committees/ institutional review boards in each country. The first study participants were enrolled on 7-June-2019 for IB1001-202 (GM2); 4-Sep-2019 for IB1001-201 (NPC); 09-Jan-2019 for IB1001-203 (A-T).

## Data Availability

Not applicable as the study is a trial design and data will be published after the trials are complete.

## List of abbreviations

8MWT: 8-meter walk test
9HPT-D: 9-hole peg test of the dominant hand
A-T: Ataxia Telangiectasia
CI-CS: clinical impression of change in severity
CI-S: clinical impression of severity
DSMB: Data Safety Monitoring Board
EQ-QoL: Euro Quality of Life
EP: Extension Phase
FDA: Food and Drug Administration
GM2: GM2 Gangliosidoses
mITT: modified intention to treat
mDRS: Modified Disability Rating Scale
NPC: Niemann-Pick type C
NPC-CSS: NPC clinical severity scale
PI: principal investigator
SARA: Scale for the Assessment and Rating of Ataxia
SCAFI: Spinocerebellar Ataxia Functional Index.

## Declarations

N/A

## Ethics approval and consent to participate

The studies have been approved by the responsible ethics committees / institutional review boards, as well as national authorities in Germany, Slovakia, Spain, United Kingdom, United States.

## Consent for publication

Not applicable.

## Availability of data and materials

Data sharing is not applicable to this article as no datasets were generated or analyzed during the current studies (study protocols).

## Authors’ contributions

MS conceived the initial studies. MS, TB initiated the initial case series, and FP, AG, GC, MS initiated the non-clinical studies, together establishing the current study concept and rational.

All authors made contributions to the conception and/ or the trial design.

TF, RKay conceptualized the novel primary outcome assessment, and MP, MS, IB, WD, WE, JG, DP, RK, DL, TM, and MF made substantial contributions to the design of the CI-CS.

TF and MS wrote and revised the study protocol. MP, TB, IB, UG, RK, TMeyer, LT made contributions to the study protocol and revised the final protocol for content. RKay has the main responsibility for statistical analyses and made substantial contributions to the study protocol. GB had the main responsibility for medical monitoring and pharmacovigilance oversight and made substantial contributions to the study protocol. CV and SF had the main responsibility for study conduct and made substantial contributions to the study protocol.

TF, MF, UG, TMeyer, CV, and SF contributed decisively to ethics approval to ethics and regulatory authorities.

MF is the sponsor’s delegated person. MS is the sponsor’s delegated clinician. TF, SF, CV are responsible for the implementation of the study.

TF and MS wrote and revised the manuscript. RKay developed the statistical concept and performed the sample size calculation. MP, GB, IB, FP, WE revised the final manuscript for content.

All authors read and approved the final manuscript.

## Acknowledgements

We thank each of the study sites and Principal Investigators participating in the IB1001 clinical trials for their participation outstanding conduct of the studies. We thank the multinational Patient Organizations representing the NPC, GM2, A-T, and related rare-disease communities, for their essential involvement, including: *NPC Organizations:* Australian Niemann-Pick Type C Foundation, Fundación Niemann-Pick de España (Spain), International Niemann-Pick disease Alliance (Worldwide), National Niemann-Pick Disease Foundation (USA), Niemann-Pick Association of Fuenlabrada (Spain), Niemann-Pick Selbsthilfegruppe (Germany), Niemann-Pick Suisse (Switzerland), Niemann-Pick UK (UK). *GM2 Organizations:* Acción y Cura para Tay-Sachs (Spain), Associação Nacional D O C E T (Portugal), Cure and Action for Tay-Sachs Foundation (UK), Cure Tay-Sachs Foundation (USA), Hand in Hand gegen Tay-Sachs und Sandhoff (Germany), National Tay-Sachs and Allied Disease Association (USA), Vaincre les Maladies Lysosomales (France). *Ataxia Organizations:* Asociación Española Familia Ataxia-Telangiectasia (Spain) Associazione Nazionale A-T (Italy), A-T Children’s Project (USA), Ataxia-Telangiectasia Society (UK), Ataxia UK (UK), ATEurope (EU), National Ataxia Foundation (USA). We thank the National Regulatory Agencies, including the Dutch Medicines Evaluation Board, German Federal Institute for Drugs and Medical Devices, Spanish Agency of Medicines and Medical Products, Slovak Human and Veterinary Medicines Agencies, US Food and Drug Administration, and UK Medicines and Healthcare products Regulatory Agency for their critical collaboration. Without their considerable effort to understand the unique challenges of drug development for NPC, GM2, and A-T patients, and actively help define a development program which expedites the availability of promising treatments for these patients, the IB1001 development program would not be feasible. Finally, we are very grateful to all the patients and their families who participated in these studies.

## Authors’ information (optional)

M. Strupp is Joint Chief Editor of the Journal of Neurology, Editor in Chief of Frontiers of Neuro-otology and Section Editor of F1000.

## Supplementary information

### Supplementary Material I: Video Acquisition, Submission, and Review

The Medpace Core Laboratory is contracted to manage the collection, quality control, central review, and storage of the subject video data acquired for the IB1001 studies. Medpace Core Laboratory provides a video acquisition manual to each clinical site, as well as a GoPro 7 (recorder), a kit of camera accessories, and a tripod. All videos should be recorded using the equipment and according to the acquisition manual provided by Medpace Core Laboratory to ensure consistency and minimize variability.

There are no specific qualifications for the designated clinical site videographer(s). Any individual at the site willing to undergo video acquisition, export, and upload training will be eligible. Each potential videographer is asked to submit two qualification “sample” videos for the purpose of qualification. The videos should be of a volunteer performing the 8MWT and 9HPT-D, acquired using the instructions outlined in a video acquisition manual. Informed consent from the volunteer(s) is obtained (if applicable). The qualification sample video submissions will confirm the ability of the clinical site to acquire and export videos in the correct file format and electronically submit to Medpace Core Laboratory.

Video informed consent from each patient will be acquired. Videos will be collected by the clinical sites and submitted to Medpace Core Laboratory for collection, quality control, central review, and storage. All patient videos should be performed by the same videographer if possible, to ensure consistency and reduce the potential for variability. All study-related video files are submitted to Medpace Core Laboratory via the web-based ClinTrak Imaging Windows Client Application. All submitted videos undergo QC (of the submitted transmittal form and storage in the ClinTrak Imaging system) within two business days of receipt by Medpace Core Laboratory. If a video is of unacceptable quality during the quality assessment, Medpace Core Laboratory may request a repeat video which should be performed as soon as possible, assuming this is feasible as determined by the investigator. Even if the video cannot be repeated, the purpose of the quality assessment is to identify issues and prevent repetition of similar quality issues on subsequent videos.

### Supplementary Material II Safety Parameters

#### Laboratory examinations - blood

A routine blood sample will be taken to exclude liver or kidney failure, and a pregnancy test for women of childbearing potential will be performed. The following laboratory parameters will be assessed at each visit (except Visit 0) via blood draw: hemoglobin, erythrocytes, hematocrit, thrombocytes, leukocytes, sodium, lactate dehydrogenase, potassium, creatinine, serum bilirubin level, aspartate aminotransferase, alanine aminotransferase, urea, alkaline phosphatase, follicle-stimulating hormone (for postmenopausal women only). Rum bicarbonate, chloride, calcium and phosphorus will also be assessed at each visit (except Visit 0) for the IB1001-203 study.

Sparse Pharmacokinetic blood sampling will be conducted during the initial treatment sequence (Visit 1 to Visit 6) at every visit. Blood samples for the quantification of N-acetyl-L-leucine in plasma will be obtained at Visit 7 and Visit 9. The first sample will be taken before first /last dosing of N-acetyl-L-leucine in the Extension Phase; subsequent samples will be taken at 30 minutes (+/- 5 minutes), 60 minutes (+/- 5 minutes), 90 minutes (+/- 10 minutes), 120 minutes (+/- 10 minutes), 150 (+/- 15 minutes), 180 minutes (+/- 15 minutes), 240 minutes (+/- 15 minutes), and 360 minutes (+/- 15 minutes) after the first/last extension phase IB1001 dose.

The total amount of blood taken per subject during the first treatment sequence (Visit 1 – Visit 6) will be 66 mL (42 mL blood for the safety analyses, 24 mL blood for the Pharmacokinetic analyses). The total amount of blood taken per subject during the Extension phase treatment sequence is 85 mL – 92 mL (21 mL – 28 mL (depending on the date of Visit 7) for safety analysis, and 64 mL for the Pharmacokinetic analysis.

#### Laboratory examinations – urine

The following laboratory parameters will be assessed via urine at each visit (except Visit 0): Leukocytes, Nitrite, Urobilinogen, Protein, pH, Occult blood (erythrocytes, leucocytes), Specific gravity, Ketones, Bilirubin, Glucose.

Urine samples will also be collected for concentrations of N-acetyl-D-leucine at the time points designated on the schedule of events. At Visit 1, this Urine sample score serves as a key enrollment criterion testing for the use of the prohibited medication N-acetyl-DL-leucine.

#### Height, Weight, Vital Signs, Physical Exam

Patient’s height, weight, systolic/diastolic blood pressure and pulse will be measured routinely throughout the duration of the study according to the schedule of events (Suppl. Table 1 and 2).

A complete physical examination that is limited to the following body systems will be conducted according to the schedule of events (Suppl. Table 1 and 2): general appearance (including skin), head and neck, eyes and ears, nose and throat, pulmonary, cardiovascular, gastrointestinal, musculoskeletal, lymphatic, and neurological.

#### Electrocardiograms

12-lead electrocardiograms will be performed at the time points designated on the schedule of events (Suppl. Table 1 and 2) and include the following measurements: Corrected QT, and whether the electrocardiograms is Normal, Abnormal not clinically significant, or Abnormal clinically significant for the patient. Intervals of the electrocardiograms will be recorded in the electronic case report form. Electrocardiograms may be performed at other visits at the discretion of the Investigator.

#### Concomitant drug and non-drug therapies

Trial participants should not begin physiotherapy or speech therapy while they are enrolled in the trial. If they are already under therapy, the number of sessions of physiotherapy and speech therapy (measured in hours of therapy per week) will be documented in the patient’s medical record and in the electronic case report form during the trial and for 6 weeks prior to screening. To adhere to the protocol, the non-pharmacological concomitant therapy should be consistent 6-weeks before Visit 1 and continued with the same intensity while the patient is enrolled in the trial.

Guided by the eligibility criteria, the administration of aminopyridines, Riluzole, Varenicline, Gabapentin or Chlorzoxazone is not allowed during the trial because of a possible beneficial effect in Cerebellar Ataxia. Variants of the IMP, -Acetyl-DL-Leucine (e.g. Tanganil^®^) or N-acetyl-L-leucine (if not provided as IMP) are not allowed during the trial. Sulfasalazine and Rosuvastatin are not allowed because drug interactions with substrates of transporters MDR1, BCRP, or BSEP cannot be excluded as N-acetyl-L-leucine is an inhibitor of these transporters *in vitro*.

#### Participation discontinuation

Patients may withdraw from the study at any time at their own request without stating the reason(s) for withdrawal. The Principal Investigator may decide that a patient should be withdrawn from the study or from the study drug. Patients who discontinue study drug prior to completing the full treatment period will be asked to complete the remaining study visits as far as possible and complete safety assessments at a minimum. If unwilling to complete the remaining study visits, regardless of the reason for withdrawal, best efforts should be made to have the patient take part in early termination procedures, preferably 7-14 days following the last dose of the study drug, unless the patient is lost to follow-up or has withdrawn his/her consent to further study participation. The reason for withdrawal (if available) will be recorded in the electronic case report form.

#### Safety assessment

The Principal Investigator is responsible for monitoring the safety of patients who have been enrolled in this study and for accurately documenting and reporting information as described in this section. In addition, the investigator will monitor the degree of stress to patients and the risk threshold throughout the trial.

Patients will be instructed to report to the Principal Investigator any Adverse Events that they experience. The Principal Investigator will ask about the occurrence of Adverse Events at each visit. Investigators are required to document all Adverse Events occurring during the clinical study, commencing with the signing of the informed consent form through the End of Study Visit (scheduled at 42 days post last IB1001 dose). Adverse event recording will continue for patients who discontinue study treatment but remain on-study, until their early termination Visit has been completed. The Principal Investigator will judge the intensity (mild, moderate, or severe), seriousness, and causality (not related or related) of all adverse events.

All adverse events will be listed by trial site and patient and displayed in summary tables that provide data combined across all sites. The incidence of adverse events and their relationship to the study drug will be analyzed descriptively, guided by the Medical Dictionary for Regulatory Activities classification.

### Supplementary Material III: Data collection

Study data for all patients will be collected in a confidential fashion using an electronic case report form supported by Viedoc. Access to the electronic case report form is restricted to staff members not involved in any aspect of the blinded evaluations. All the information required by the protocol must be documented and any omissions explained. The Investigator must review all electronic case report form entries for completeness and accuracy. Source documents, including all demographic and medical information, electronic case report form and informed consent form for each patient in the study must be maintained by the Investigator. All information in the electronic case report form must be traceable to the original source documents. An audit trail of all changes to this database, including the date, reason for the data change and who made the change, will be maintained within the same database. The audit trail will be part of the archived data at the end of the study. Concomitant medication and adverse events will be coded using standardized medical dictionaries.

**Supplementary Table 1.** Parent Study schedule of enrolment, interventions, and assessments.

| Period in the Parent Study | Study Run-In |  | Baseline Period |  | Treatment Period |  | Wash-Out Period |  | Early Term. |
| --- | --- | --- | --- | --- | --- | --- | --- | --- | --- |
|  | Screening visit | Pre-treatment Washout |  |  |  |  |  |  |  |
| Duration of the whole period | 1 Day | 6 Weeks | 2 Weeks |  | 6 Weeks |  | 6 Weeks |  | 1 Day |
| Visit number | Visit 0 |  | Visit 1 | Visit 2 | Visit 3 | Visit 4 | Visit 5 | Visit 6 / End Parent Study | Early Term |
| Name of the Visit | Screening |  | Baseline 1 | Baseline 2/ Start IB1001 | Treatment 1 | Treatment 2 | Washout 1 | Washout 2 | Early Term |
| Timeline (Days) | Day -56 |  | Day -14 | Day 1 | Day 28 | Day 42 | Day 70 | Day 84 | na |
| Visit Window allowed | na |  | na | +7 days | +7 days | +7 days | +7 days | +7 days | na |

**Supplementary Table 1**
|  | Non-naïve patients only | Non-naïve patients only |  |  |  |  |  |  |  |
| --- | --- | --- | --- | --- | --- | --- | --- | --- | --- |
| Patient information and informed consent process | X |  | X<sup>1</sup> |  |  |  |  |  |  |
| Inclusion / exclusion criteria | X |  | X | X |  |  |  |  |  |
| Patient weight and height measurements | X |  | X<sup>1</sup> |  |  |  |  |  |  |
| Physical Exam (203 study only) |  |  |  |  |  |  |  |  |  |
| Vital signs | X |  | X | X | X | X | X | X | X |
| Patient demographics (in accordance with local regulations) | X |  | X<sup>1</sup> |  |  |  |  |  |  |
| Relevant medical history | X |  | X<sup>1</sup> |  |  |  |  |  |  |
| 60-Day drug history | X |  | X<sup>1</sup> |  |  |  |  |  |  |
| Documentation of concomitant medication | X |  | X | X | X | X | X | X | X |
| Documentation of therapy | X |  | X | X | X | X | X | X | X |
| Confirmation prohibited medications have not been used in the past 42 days at Visit 1/since Visit 1 for Visit 2 |  |  | X | X |  |  |  |  |  |
| Confirmation prohibited medications have been used within past 42 days | X |  |  |  |  |  |  |  |  |
| Classify patient as “Naïve” or “Non-naïve” | X |  | X<sup>1</sup> |  |  |  |  |  |  |
| 12-lead electrocardiogram |  |  | X |  | X |  | X |  | X |

|  | Non-naïve patients only | Non-naïve patients only |  |  |  |  |  |  |  |
| --- | --- | --- | --- | --- | --- | --- | --- | --- | --- |
| Urine Test for N-Acetyl-D-Leucine |  |  | X | X |  |  | X | X | X |
| Urine by dipstick for pregnancy test (if applicable) |  |  | X<sup>2</sup> | X |  | X |  | X | X |
| Urine by dipstick for pregnancy test (if applicable) |  |  | X<sup>2</sup> | X |  | X |  | X | X |
| Urinalysis |  |  | X | X | X | X | X | X | X |
| Blood safety laboratory tests |  |  | X | X | X | X | X | X | X |
| Blood sample for sparse PK |  |  | X | X | X | X | X | X | X |
| Quality of Life EQ-5D-5L for patients aged ≥18 years; EQ-5D-Y for children aged <18 years |  |  | X | X | X | X | X | X | X |
| Scale for Ataxia Rating (SARA) | X |  | X | X | X | X | X | X | X |
| Scale for Spinocerebellar Ataxia Functional Index (SCAFI) | X |  | X | X | X | X | X | X | X |
| Cognitive assessment according to standard procedures of the clinical site |  |  | X |  |  |  |  |  |  |
| Determination of CI-CS Primary Anchor Test (9HPT-D or 8MWT) |  |  | X |  |  |  |  |  |  |
| Modified Disabling Rating Score (201 and 202 study only) |  |  | X | X | X | X | X | X | X |
| Niemann-Pick Disease type C Clinical Severity Scale (201 study only) |  |  | X |  |  |  |  |  |  |
| Columbia Suicide Severity Rating Scale (203 study only) | X |  | X | X | X | X | X | X | X |

|  | Non-naïve patients only | Non-naïve patients only |  |  |  |  |  |  |  |
| --- | --- | --- | --- | --- | --- | --- | --- | --- | --- |
| Clinical Global Impression of Severity by Physician, Caregiver, Patient (if able) |  |  | X | X | X | X | X | X | X |
| Clinical Global Impression of Improvement by Physician, Caregiver, Patient (if able) |  |  |  |  |  | X |  | X | X |
| Documentation of adverse events | X |  | X | X | X | X | X | X | X |
| Dispensing of study drug |  |  |  | X | X |  |  |  |  |
| Intake of study drug at site |  |  |  | X |  |  |  |  |  |
| Return of study drug |  |  |  |  | X | X |  |  | X |
| Study drug compliance check |  |  |  |  | X | X |  |  | X |

<sup>1</sup> "Naïve" patient only
<sup>2</sup> IB1001-203 only

**Supplementary Table 2.**
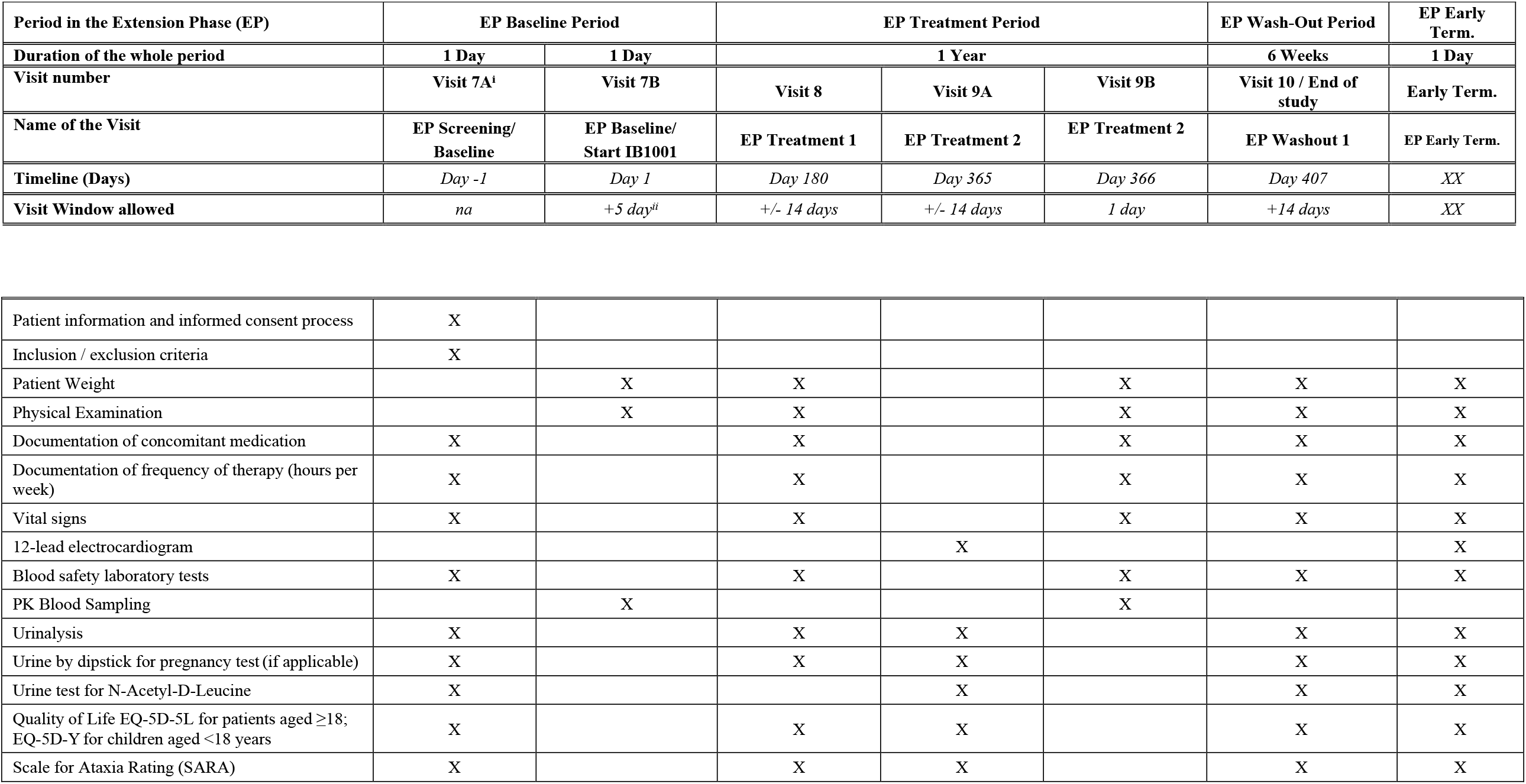
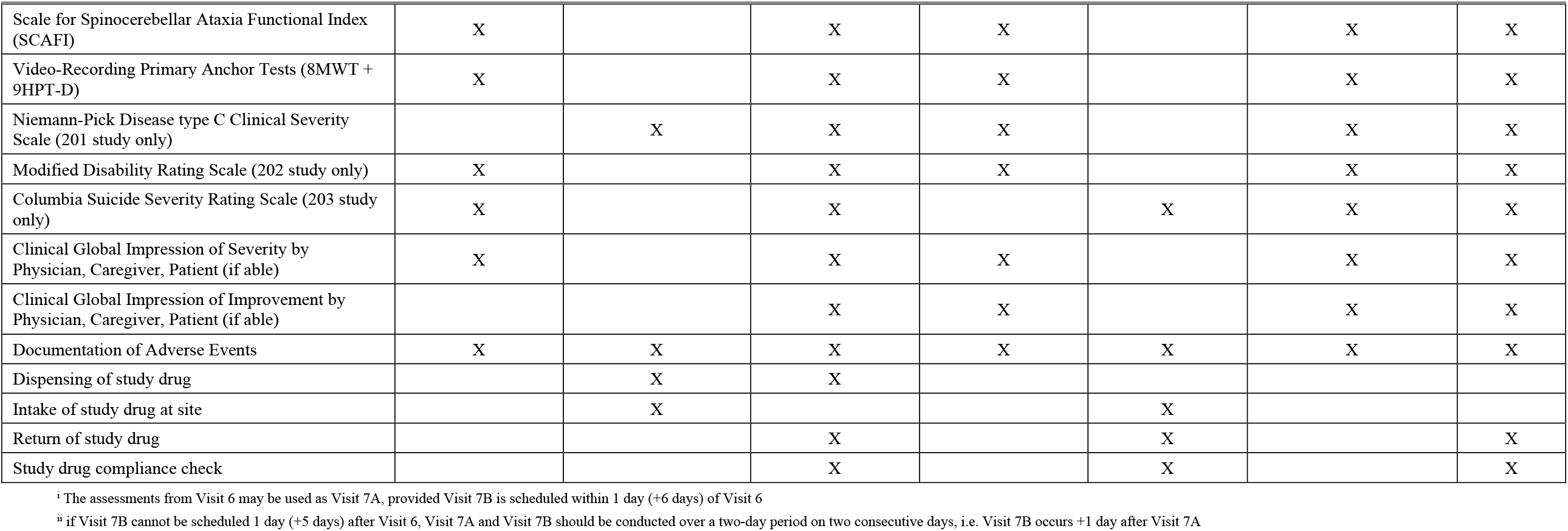
Extension Phase schedule of enrolment, interventions, and assessments.

| Period in the Extension Phase (EP) | EP Baseline Period |  | EP Treatment Period |  |  | EP Wash-Out Period | EP Early Term. |
| --- | --- | --- | --- | --- | --- | --- | --- |
| Duration of the whole period | 1 Day | 1 Day | 1 Year |  |  | 6 Weeks | 1 Day |
| Visit number | Visit 7A<sup>i</sup> | Visit 7B | Visit 8 | Visit 9A | Visit 9B | Visit 10 / End of study | Early Term. |
| Name of the Visit | EP Screening/<br>Baseline | EP Baseline/<br>Start IB1001 | EP Treatment 1 | EP Treatment 2 | EP Treatment 2 | EP Washout 1 | EP Early Term. |
| Timeline (Days) | Day -1 | Day 1 | Day 180 | Day 365 | Day 366 | Day 407 | XX |
| Visit Window allowed | na | +5 day<sup>ii</sup> | +/- 14 days | +/- 14 days | 1 day | +14 days | XX |

| Supplementary Table 2 |  |  |  |  |  |  |  |
| --- | --- | --- | --- | --- | --- | --- | --- |
| Patient information and informed consent process | X |  |  |  |  |  |  |
| Inclusion / exclusion criteria | X |  |  |  |  |  |  |
| Patient Weight |  | X | X |  | X | X | X |
| Physical Examination |  | X | X |  | X | X | X |
| Documentation of concomitant medication | X |  | X |  | X | X | X |
| Documentation of frequency of therapy (hours per week) | X |  | X |  | X | X | X |
| Vital signs | X |  | X |  | X | X | X |
| 12-lead electrocardiogram |  |  |  | X |  |  | X |
| Blood safety laboratory tests | X |  | X |  | X | X | X |
| PK Blood Sampling |  | X |  |  | X |  |  |
| Urinalysis | X |  | X | X |  | X | X |
| Urine by dipstick for pregnancy test (if applicable) | X |  | X | X |  | X | X |
| Urine test for N-Acetyl-D-Leucine | X |  |  | X |  | X | X |
| Quality of Life EQ-5D-5L for patients aged ≥18; EQ-5D-Y for children aged <18 years | X |  | X | X |  | X | X |
| Scale for Ataxia Rating (SARA) | X |  | X | X |  | X | X |
| Scale for Spinocerebellar Ataxia Functional Index (SCAFI) | X |  | X | X |  | X | X |
| Video-Recording Primary Anchor Tests (8MWT + 9HPT-D) | X |  | X | X |  | X | X |
| Niemann-Pick Disease type C Clinical Severity Scale (201 study only) |  | X | X | X |  | X | X |
| Modified Disability Rating Scale (202 study only) | X |  | X | X |  | X | X |
| Columbia Suicide Severity Rating Scale (203 study only) | X |  | X |  | X | X | X |
| Clinical Global Impression of Severity by Physician, Caregiver, Patient (if able) | X |  | X | X |  | X | X |
| Clinical Global Impression of Improvement by Physician, Caregiver, Patient (if able) |  |  | X | X |  | X | X |
| Documentation of Adverse Events | X | X | X | X | X | X | X |
| Dispensing of study drug |  | X | X |  |  |  |  |
| Intake of study drug at site |  | X |  |  | X |  |  |
| Return of study drug |  |  | X |  | X |  | X |
| Study drug compliance check |  |  | X |  | X |  | X |

<sup>i</sup> The assessments from Visit 6 may be used as Visit 7A, provided Visit 7B is scheduled within 1 day (+6 days) of Visit 6
<sup>ii</sup> if Visit 7B cannot be scheduled 1 day (+5 days) after Visit 6, Visit 7A and Visit 7B should be conducted over a two-day period on two consecutive days, i.e. Visit 7B occurs +1 day after Visit 7A

